# Natriuretic Peptides Improve Classification of People at Low Risk of Atrial Fibrillation after Stroke

**DOI:** 10.1101/2023.09.28.23296282

**Authors:** Alan Cameron, Markus Arnold, Georgios Katsas, Jason Yang, Terence J Quinn, Azmil H Abdul-Rahim, Ross Campbell, Kieran Docherty, Gian Marco De Marchis, Marcel Arnold, Timo Kahles, Krassen Nedeltchev, Carlo Cereda, Georg Kaegi, Alejandro Bustamante, Joan Montaner, George Ntaios, Christian Foerch, Katharina Spanaus, Arnold Von Eckardstein, Jesse Dawson, Mira Katan

## Abstract

**Background:** Prolonged cardiac monitoring (PCM) increases atrial fibrillation detection after stroke (AFDAS) but access is limited. We aimed to assess the utility of midregional pro-atrial natriuretic peptide (MR-proANP) and N-terminal pro-B-type natriuretic peptide (NT-proBNP) to identify people who are unlikely to have AFDAS and improve healthcare resource allocation for PCM..

**Methods:** We analysed people from the BIOSIGNAL (Biomarker Signature of Stroke Aetiology) study with ischaemic stroke, no known AF and ≥3 days cardiac monitoring. External validation was in the PRECISE (Preventing Recurrent Cardioembolic Stroke: Right Approach, Right Patient) study of 28-days cardiac monitoring after stroke.

The main outcome is no AFDAS. We assessed the discriminatory value of MR-proANP and NT-proBNP combined with clinical variables to identify people with no AFDAS. We determined the net reduction in people who would undergo PCM using the models with 15% AFDAS threshold probability.

**Results:** We included 621 people from BIOSIGNAL. The clinical model included age, National Institutes of Health Stroke Scale score, lipid-lowering therapy, creatinine and smoking status. The AUROC was 0.68 (95%CI 0.62-0.74) with clinical variables, which improved with log_10_MR-proANP (0.72,0.66-0.78;p=0.001) or log_10_NT-proBNP (0.71,0.65-0.77;p=0.009). Performance was similar for log_10_MR-proANP versus log_10_NT-proBNP (p=0.28).

In 239 people from PRECISE, the AUROC for clinical variables was 0.68 (0.59-0.76), which improved with log_10_NT-proBNP (0.73,0.65-0.82;p<0.001) or log_10_MR-proANP (0.79,0.72-0.86;p<0.001). Performance was better with log_10_MR-proANP versus log_10_NT-proBNP (p=0.03).

The models could reduce the number who would undergo PCM by 30% (clinical+log_10_MR-proANP), 27% (clinical+log_10_NT-proBNP) or 20% (clinical).

**Conclusions:** MR-proANP and NT-proBNP help classify people who are unlikely to have AFDAS and could reduce the number who need PCM by 30%.

## Introduction

Atrial fibrillation (AF) accounts for 25% of ischaemic strokes.^1^ One third of strokes are recurrent and anticoagulant drugs reduce the risk of recurrent strokes in people with AF.^2,3^ People are screened for AF detection after stroke (AFDAS) or transient ischaemic attack (TIA) with a cardiac monitor. Typically, this has two stages: 1) an initial period of cardiac monitoring in all people after stroke; and 2) longer-term monitoring in people with unknown stroke aetiology. For the first stage, standard practice is an electrocardiogram (ECG) and a 24-72 hour Holter monitor that detects AF in ∼4% of people.^4,5^

Prolonged cardiac monitoring (PCM) identifies more cases of AFDAS.^6–9^ An external loop recorder for 30 days detects AF in 16% of people with ischaemic stroke of unknown aetiology.^6^ An implantable loop recorder (ILR) for one year can identify AF in up to 24% of such people.^7^ National/international guidelines recommend PCM to search for AFDAS, especially in people with ischaemic stroke or TIA of unknown aetiology.^10,11^ However, PCM is costly and is not widely available.^12^ Further, while the yield of AFDAS is higher with PCM, most people tested do not have AF.^6,7^

This could be addressed by better patient selection. A solution could be to identify people who are unlikely to have AFDAS and who do not need PCM. Prognostic scores exist for AFDAS, although these focus on people with embolic stroke of undetermined source (ESUS) who have undergone a period of monitoring or identifying people who are most likely to have AFDAS.^13–17^ An alternative approach is to identify people who are unlikely to have AFDAS and “rule out” rather than “rule in”.^18^ This would allow PCM to be focused on people who are more likely to have AFDAS and benefit from anticoagulation to prevent second AF-related strokes, while minimising potentially unnecessary tests for people who are unlikely to have AFDAS.

The natriuretic peptides midregional pro-atrial natriuretic peptide (MR-proANP) and N-terminal pro-B-type natriuretic peptide (NT-proBNP) could help stratify probability of AFDAS.^19–21^

### Study aim

We aimed to assess the utility of MR-proANP and NT-proBNP to identify people who are unlikely to have AFDAS and improve healthcare resource allocation for PCM.

## Methods

### Study Design and Patient Population

We analysed people in the Biomarker Signature of Stroke Aetiology Study (BIOSIGNAL) study (NCT02274727) who were not known to have AF and who had ≥3 days cardiac monitoring (Supplemental Figure 1). The BIOSIGNAL study was a multi-centre, prospective cohort study that evaluated blood biomarkers associated with underlying stroke aetiology in people after ischaemic stroke.^20^ People with ischaemic stroke were recruited from nine European stroke centres between October 2014 and October 2017.

### Study Assessments and Intervention

Brain imaging, assessment of demographic variables and biochemical profile was performed on admission.

#### Biomarker measurement

Blood samples were collected within 24 hours of symptom onset and separated by centrifugation at 3,000g and 4°C for 20 minutes. Plasma samples were aliquoted and frozen at -80°C. MR-proANP concentrations (pmol/L) were measured using automated B.R.A.H.M.S MR-proANP immunoassay technology on a KRYPTOR instrument (B.R.A.H.M.S GmbH).^22^ Plasma was analysed for NT-proBNP concentrations (ng/l) using a Cobas 8000 analyser (Roche Diagnostics, Basel, Switzerland).

#### Cardiac rhythm monitoring

Participants had an ECG and 24 hours cardiac monitoring on admission. People with no AFDAS in the first 24 hours received further monitoring according to standard operating procedures of enrolling centres, ideally for ≥48 hours.^4^ In people who still did not have evidence of AFDAS, further monitoring was recommended, preferably with a 7-day ECG monitor or implantable cardiac device. Follow-up was at 3 months and 12 months during an outpatient visit or telephone interview with a stroke physician.

### Outcome measures

The main outcome is no AFDAS. AF was defined as ≥30 seconds of AF/atrial flutter, based on international guidelines that define clinical AF as an episode ≥30 seconds^4^ and is recommended as an indication for anticoagulation.^23^ Central adjudication of outcomes was conducted by certified vascular neurologists and trained neurovascular fellows. In cases of disagreement, consensus was reached after discussion. To minimize bias, all outcome evaluators were blinded to biomarker levels.

### Statistical analyses

#### Demographics

Discrete variables are expressed as counts (%) and continuous variables as median (IQR). Baseline demographics and risk factors were compared using t-tests or a non-parametric equivalent. Logarithmic transformation (log_10_) was used to normalize skewed distributions.

#### Analyses

Univariable and multivariable logistic regression analyses were performed to evaluate associations with demographic variables and no AFDAS. Area under receiver-operating-characteristic (AUROC) curves were constructed incorporating clinical variables associated with AFDAS in univariable analysis (age, NIHSS score, creatinine, smoking and lipid-lowering medication) and log_10_MR-proANP or log_10_NT-proBNP using the pROC package and 95% confidence intervals were derived using bootstrap sampling.^24^ We compared AUROCs for models with clinical variables versus clinical variables and log_10_MR-proANP or log_10_NT-proBNP using the Likelihood-Ratio Test for nested models. We compared AUROCs for models with clinical variables and log_10_MR-proANP versus log_10_NT-proBNP using De Long’s test and continuous Net Reclassification Index (NRI).^25^ All statistical analyses were performed using R version 4.3.

#### Sensitivity Analyses

Sensitivity analyses were conducted in subgroups, including: people older than the median age; female gender; NIHSS score > median; and cardiac disease, defined as prior myocardial infarction (MI) or heart failure. Sensitivity analyses were also conducted in people from the BIOSIGNAL study with i) any duration of cardiac monitoring; and ii) <3 days cardiac monitoring.

### External Validation

External validation of the models with clinical variables ± log_10_MR-proANP or log_10_NT-proBNP was performed in people recruited to the ongoing Preventing Recurrent Cardioembolic Stroke: Right Approach, Right Patient (PRECISE) study.^26^ The PRECISE study is a prospective cohort study of 28 days cardiac monitoring in people with ischaemic stroke or TIA who are not known to have AF. Participants are recruited from two Scottish stroke centres since 2021. Venous blood is collected within 5 days of hospital admission and separated by centrifugation prior to storage at -80°C. MR-proANP concentrations are measured using the B.R.A.H.M.S KRYPTOR instrument (B.R.A.H.M.S GmbH) and NT-proBNP concentrations are measured using a Cobas e411 analyser (Roche Diagnostics, Basel, Switzerland).

Participants receive 28 days cardiac monitoring with a Novacor R-test or ECG patch. The Novacor R-test has been used for all participants included in the current analysis. The primary outcome is AF ≥30 seconds and all AF cases are verified by two Stroke Physicians and two Cardiologists.

### Decision Curve Analysis

We performed a decision curve analysis combining data from the BIOSIGNAL and PRECISE cohorts. We assessed the utility of the models to identify people who are unlikely to have AFDAS and reduce the number of people who would undergo PCM.^27,28^ We determined the net reduction in people who would undergo PCM with an AFDAS threshold probability of 15%, which was selected based on AF detection rates in the EMBRACE study^6^ and a meta-analysis of studies evaluating PCM after ischaemic stroke or TIA.^8^

### Ethics and regulatory approval

The BIOSIGNAL study was approved by the local Ethics Committees of participating centres (main centre: Cantonal Ethics Commission of Zurich, BASEC-Nr. PB_2016-00672) and conducted according to the Declaration of Helsinki. All participants, or their welfare guardians, provided written informed consent. The PRECISE study is approved by the West of Scotland Research Ethics Committee 3 and all participants provide written, informed consent.

## Results

### Baseline characteristics

We included 621 people from the BIOSIGNAL study: 243 (39.1%) were female, the median (IQR) age was 69 (57-78) years and 77 people (12%) were found to have AFDAS (Table 1). The median (IQR) duration of ECG monitoring was 9 (7-11) days and median (IQR) NIHSS score was 4 (2-8). MR-proANP concentrations were available for 603 participants (97.1%) and NT-proBNP levels were available for 616 participants (99.2%).

**Table 1.**
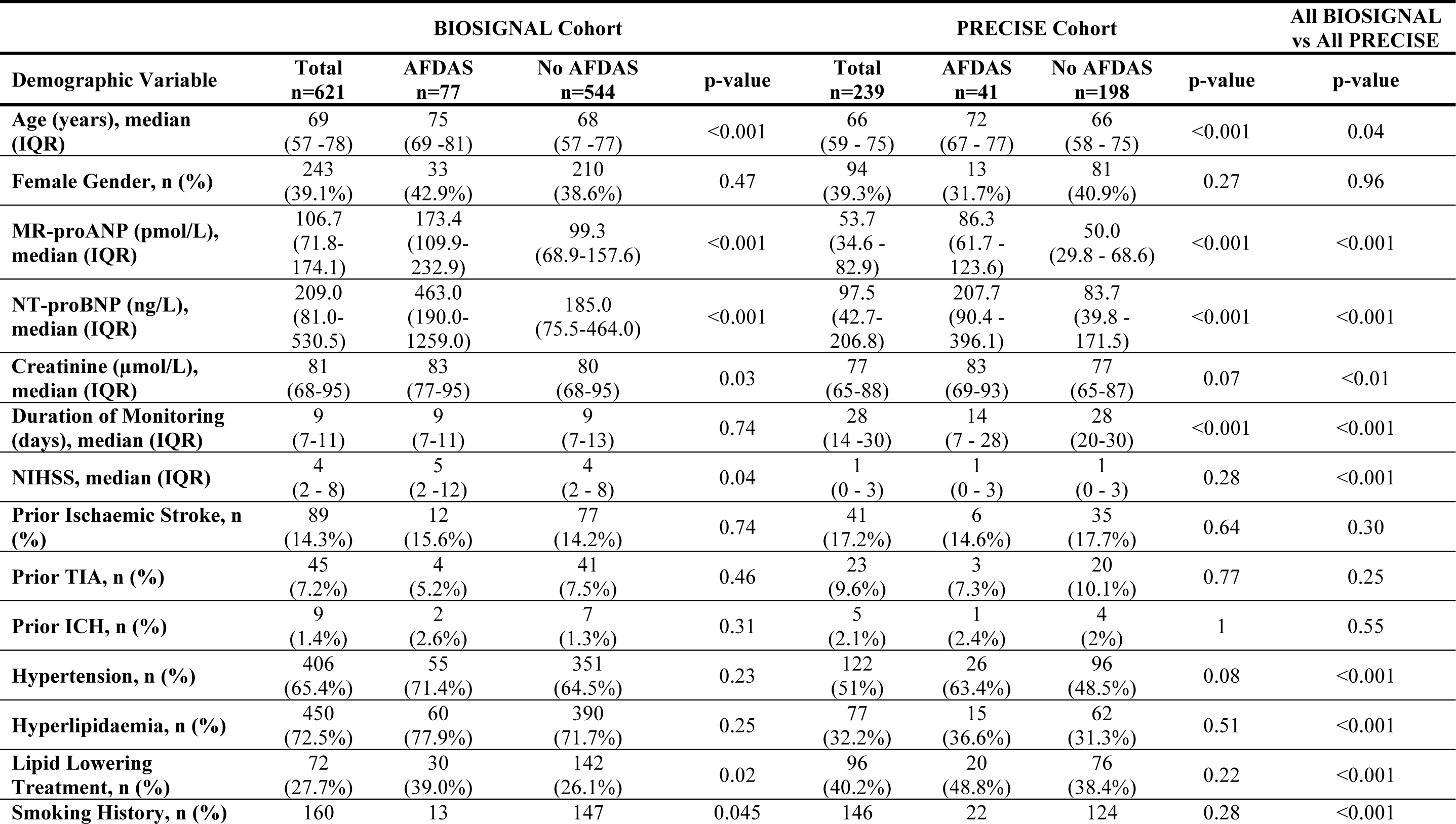
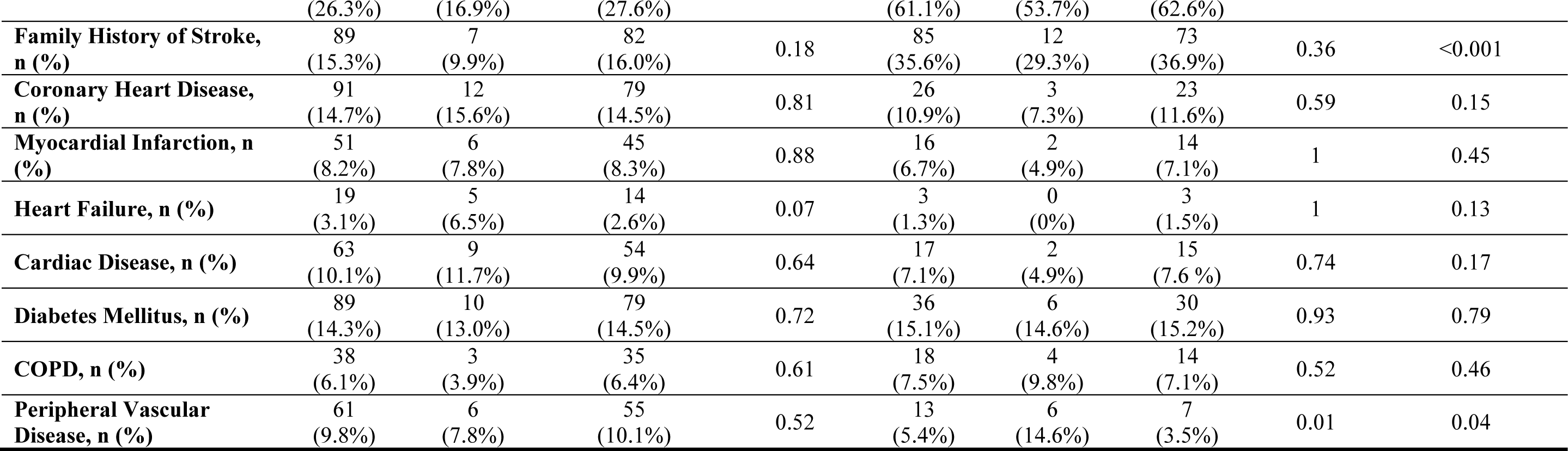
Baseline Demographics.

People with no AFDAS were younger (median (IQR) age 68 (57-77) versus 75 (69-81) years, p<0.001) and had lower MR-proANP (99.3 (68.9-157.6) versus 173.4 (109.9-232.9) pmol/L, p<0.001), NT-proBNP (185.0 (75.5-464.0) versus 463.0 (190.0-1259.0) ng/l, p<0.001) and creatinine (80 (68-95) versus 83 (77-95) µmol/L, p=0.03) (Table 1). People with no AFDAS were more likely to have a history of smoking (27.6% versus 16.9%, p=0.045) and were less likely to be prescribed lipid lowering medication at baseline (26.1% versus 39.0%, p=0.02).

### Univariable analysis

Variables associated with no AFDAS in univariable analysis were lower log_10_MR-proANP (OR 20.00, 95% CI 7.14-50.00, p<0.001), lower log_10_NT-proBNP (OR 2.78, 95% CI 1.89-4.17, p<0.001), younger age (OR 1.05, 95% CI 1.03-1.08, p<0.001), lower NIHSS score (OR 1.04, 95% CI 1.00-1.09, p=0.03), no lipid-lowering therapy (OR 1.82, 95% CI 1.10-2.94, p=0.02), lower creatinine (OR 1.01, 95% CI 1.00-1.02, p=0.02) and smoking history (OR 1.88, 95% CI 1.01-3.52, p=0.048) (Table 2).

**Table 2.**
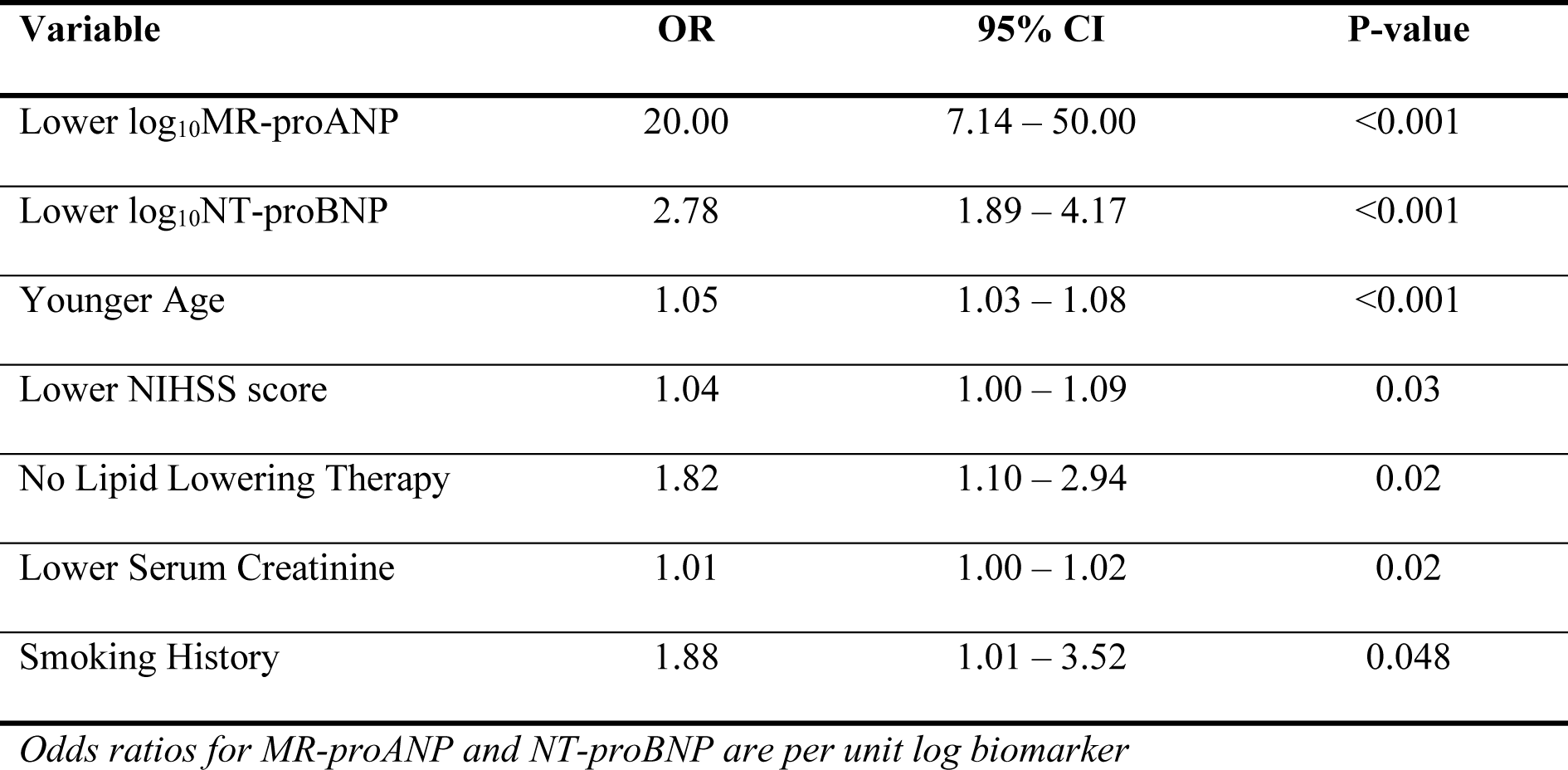
Variables Associated with No AF Detection in Univariable Analysis.

| **Variable** | **OR** | **95% CI** | **P-value** |
| --- | --- | --- | --- |
| Lower log<sub>10</sub>MR-proANP | 20.00 | 7.14 – 50.00 | <0.001 |
| Lower log<sub>10</sub>NT-proBNP | 2.78 | 1.89 – 4.17 | <0.001 |
| Younger Age | 1.05 | 1.03 – 1.08 | <0.001 |
| Lower NIHSS score | 1.04 | 1.00 – 1.09 | 0.03 |
| No Lipid Lowering Therapy | 1.82 | 1.10 – 2.94 | 0.02 |
| Lower Serum Creatinine | 1.01 | 1.00 – 1.02 | 0.02 |
| Smoking History | 1.88 | 1.01 – 3.52 | 0.048 |

*Odds ratios for MR-proANP and NT-proBNP are per unit log biomarker*

### Multivariable analysis

In multivariable analysis with log_10_MR-proANP and clinical variables associated with no AFDAS in univariable analysis, the only variable that remained associated with no AFDAS was lower log_10_MR-proANP (OR 8.33, 95% CI 2.50-25.00, p<0.01) (Table 3).

**Table 3.** Variables Associated with No AFDAS in Multivariable Analysis with log_10_MR-proANP.

| **Variable** | **OR** | **95% CI** | **P-value** |
| --- | --- | --- | --- |
| Lower log<sub>10</sub>MR-proANP | 8.33 | 2.50 – 25.00 | <0.01 |
| Younger Age | 1.02 | 1.00 – 1.05 | 0.10 |
| Lower NIHSS score | 1.02 | 0.97 – 1.06 | 0.48 |
| No Lipid Lowering Therapy | 0.99 | 0.57 – 1.72 | 0.97 |
| Lower Serum Creatinine | 1.00 | 1.00 – 1.01 | 0.45 |
| Smoking History | 1.18 | 0.60 – 2.30 | 0.63 |

*Odds ratios for MR-proANP and NT-proBNP are per unit log biomarker*

In multivariable analysis with log_10_NT-proBNP and clinical variables associated with no AFDAS in univariable analysis, the only variables that remained associated with no AFDAS were lower log_10_NT-proBNP (OR 1.85, 95% CI 1.15-2.94, p=0.01) and younger age (OR 1.03, 95% CI 1.01-1.06, p=0.01) (Table 4).

**Table 4.** Variables Associated with No AFDAS in Multivariable Analysis with log_10_NT-proBNP.

| **Variable** | **OR** | **95% CI** | **P-value** |
| --- | --- | --- | --- |
| Lower log<sub>10</sub>NT-proBNP | 1.85 | 1.15 – 2.94 | 0.01 |
| Younger Age | 1.03 | 1.01 – 1.06 | 0.01 |
| Lower NIHSS score | 1.03 | 0.99 – 1.08 | 0.16 |
| No Lipid Lowering Therapy | 1.10 | 0.65 – 1.89 | 0.72 |
| Lower Serum Creatinine | 1.00 | 1.00 – 1.01 | 0.37 |
| Smoking History | 1.31 | 0.68 – 2.53 | 0.43 |

*Odds ratios for MR-proANP and NT-proBNP are per unit log biomarker*

### AUROC analyses

The model for predicting no AFDAS with clinical variables demonstrated AUROC 0.68 (95% CI 0.62-0.74; Table 5). Performance improved when log_10_MR-proANP or log_10_NT-proBNP was added: 0.72 (95% CI 0.66-0.78) for log_10_MR-proANP (p=0.001); and 0.71 (95% CI 0.65-0.77) for log_10_NT-proBNP (p=0.009). MR-proANP and NT-proBNP performed similarly in direct comparison of models with log_10_MR-proANP versus log_10_NT-proBNP (p=0.28).

**Table 5.**
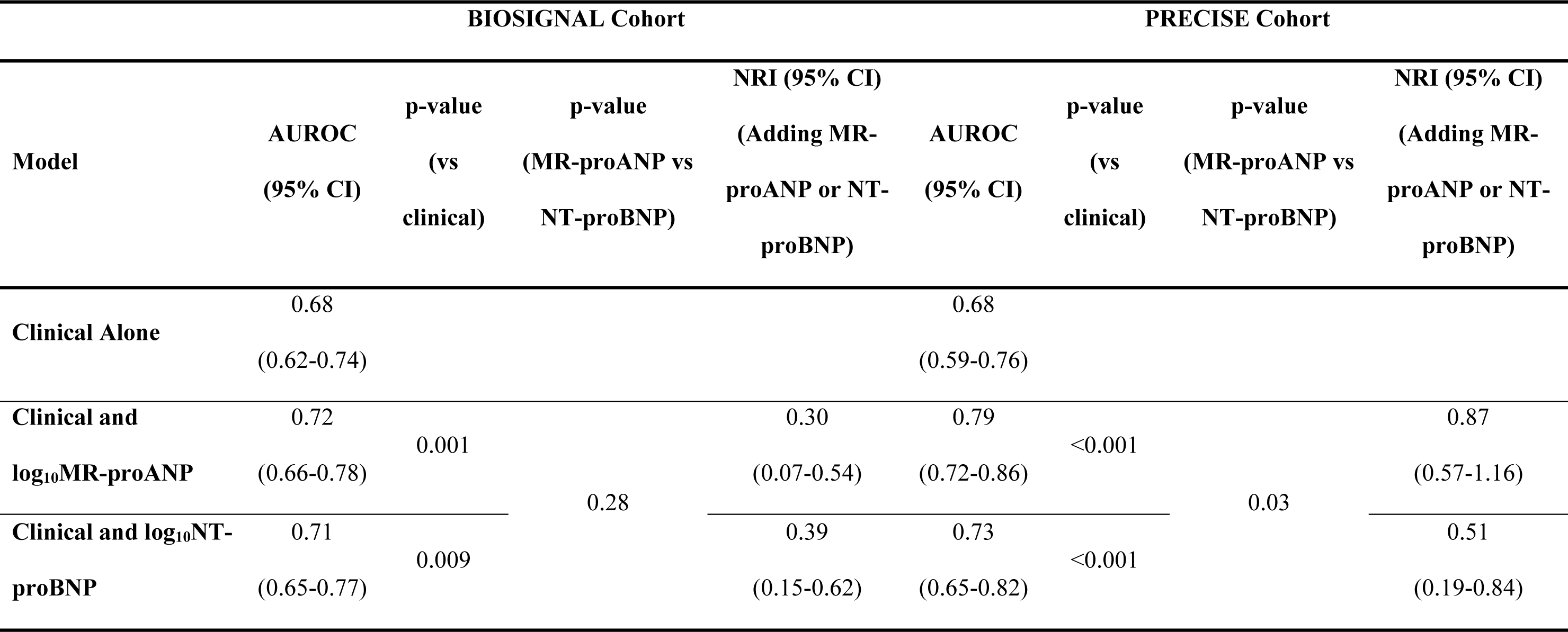
Performance of the Models with Clinical Variables ± MR-proANP or NT-proBNP in the BIOSIGNAL and PRECISE Cohorts.

Among people without AFDAS, by measuring MR-proANP a net of 55% were correctly moved towards lower probability of AFDAS, while 45% were falsely moved towards higher probability of AFDAS. The corresponding NRI was 0.30 (95% CI 0.07-0.54). Similar findings were observed for measuring NT-proBNP: a net of 55% were correctly moved towards lower probability of AFDAS, while 45% were falsely moved towards higher probability of AFDAS and the corresponding NRI was 0.39 (95% CI 0.15-0.62).

### The Impact of Baseline Demographics and Cardiac Monitoring Duration on Prediction Performance

There was no significant difference in model performance across age, gender, NIHSS score or prior cardiac disease (Supplemental Table 1). AUROCs were numerically higher in people with no prior cardiac disease for models with both natriuretic peptides. Performance of the model with clinical variables was similar in people with any duration or <3 days cardiac monitoring. AUROCs were greater for the models with log_10_MR-proANP or log_10_NT-proBNP among people with any duration or <3 days monitoring, compared to people with ≥3 days monitoring (Supplemental Table 2).

### External validation

External validation was performed in 239 people recruited to the PRECISE study: 94 (39.3%) were female, the median (IQR) age was 66 (59-75) years and the median (IQR) NIHSS score was 1 (0-3). The median (IQR) duration of cardiac monitoring was 28 (14-30) days and 41 people (17.2%) had AFDAS. MR-proANP and NT-proBNP concentrations were available for all 239 participants.

Compared to the BIOSIGNAL cohort, people in the PRECISE cohort were younger (median (IQR) age 66 (59-75) versus 69 (58-78) years, p=0.04) and had lower MR-proANP (53.7 (34.6-82.9) versus 106.7 (71.8-174.1) pmol/L, p<0.001) and NT-proBNP (97.5 (42.7-206.8) versus 209.0 (81.0-530.5) ng/L, p<0.001).

When applied to the PRECISE cohort, the AUROC was higher for the model with log_10_MR-proANP and similar for models with clinical variables or log_10_NT-proBNP (Table 5). The AUROC was 0.68 (95% CI 0.59-0.76) for clinical variables and improved with log_10_MR-proANP (0.79 (0.72-0.86), p<0.001) or log_10_NT-proBNP (0.73 (0.65-0.82), p<0.001). In the PRECISE cohort, the model with clinical variables and log_10_MR-proANP performed better than log_10_NT-proBNP (p=0.03).

Among people without AFDAS, by measuring MR-proANP a net of 68% were correctly moved towards lower probability of AFDAS, while 32% were falsely moved towards higher probability of AFDAS. The corresponding NRI was 0.87 (95% CI 0.57-1.16). Similar findings were observed for NT-proBNP: a net of 67% were correctly moved towards lower probability of AFDAS, while 33% were falsely moved towards higher probability of AFDAS and the corresponding NRI was 0.51 (95% CI 0.19-0.84).

### Clinical Utility to Reduce the Number of People who Need Prolonged Cardiac Monitoring

Applying an AFDAS threshold probability of 15%, the estimated net reduction in people who would undergo PCM is 30% for the model with log_10_MR-proANP, 27% for the model with log_10_NT-proBNP, and 20% for the model with clinical variables alone (Figure 1, Supplemental Table 3).

**Figure 1.**
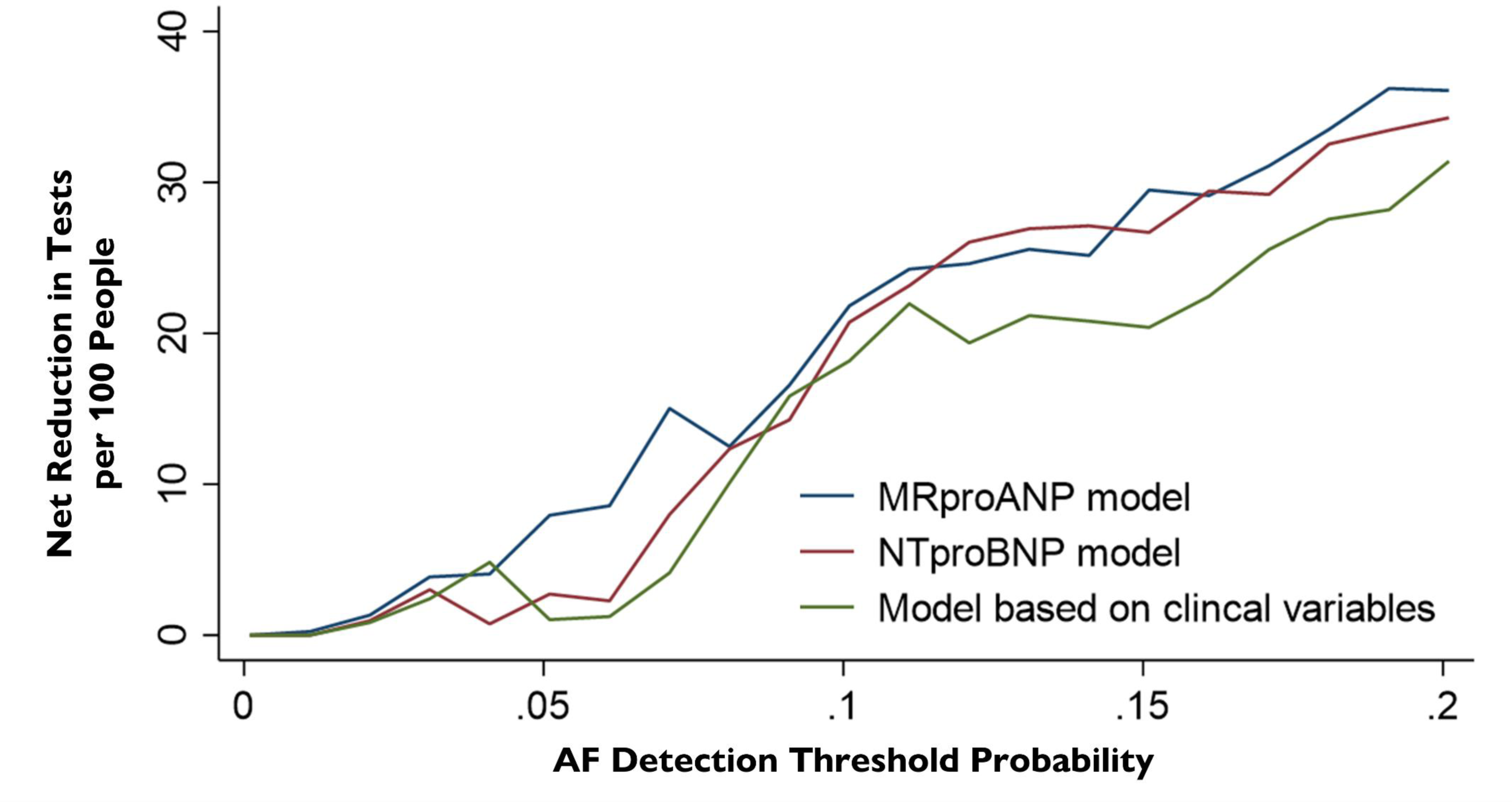
Decision Curve Analysis for Net Reduction in the Number of People who Would Need Prolonged Cardiac Monitoring.

## Discussion

Our findings, with external validation, demonstrate that MR-proANP and NT-proBNP can help identify people who are unlikely to have AFDAS and reduce the number of people who would undergo PCM by approximately 30%. Indeed, both natriuretic peptides improve performance compared to clinical variables alone. Measuring MR-proANP or NT-proBNP to guide patient selection could help focus PCM on people who are more likely to have AFDAS and benefit, while minimising testing for people who are unlikely to have AFDAS. In healthcare systems where PCM resources are limited, it is likely that such an approach would increase the proportion of people who are found to have AFDAS and who would receive anticoagulation to prevent recurrent strokes.

Our analysis is unique as we 1) focus on identifying people who are unlikely to have AFDAS (“rule out”); 2) evaluate the role of natriuretic peptide blood biomarkers; 3) include unselected people after stroke; and 4) externally validate our findings. Previous scores to stratify probability of AFDAS focus on people with ESUS who have already undergone a period of cardiac monitoring, identifying higher risk people (“rule in”) and do not include blood biomarkers.^16–18,29^

Our findings support that MR-proANP and NT-proBNP have strong potential to identify people who are unlikely to have AFDAS. Indeed, MR-proANP improved performance compared to clinical variables, and the model with MR-proANP had numerically greater AUROC and odds ratio than models with NT-proBNP or clinical variables alone. In multivariable analysis with MR-proANP, lower MR-proANP was the only variable associated with no AFDAS. In contrast, younger age and lower NT-proBNP were associated with no AFDAS in multivariable analysis with NT-proBNP.

The model with NT-proBNP was superior to clinical variables and NT-proBNP may also help identify people who are unlikely to have AFDAS. Previous studies support that NT-proBNP may help stratify probability of AFDAS. Indeed, sub-analysis of the Find-AF_RANDOMISED_ Trial demonstrates that a BNP threshold of 100 ng/L to select people for prolonged (3× 10-day) Holter ECG monitoring after stroke can reduce the number needed to screen from 18 to 3.^21^ NT-proBNP analysis is inexpensive, routinely used to screen for heart failure and may be more readily available in hospital laboratories or by using point-of-care tests than MR-proANP.^30^ Overall, both MR-proANP and NT-proBNP can help classify people who are unlikely to have AFDAS. The biomarker which performs best and would be most readily implemented in healthcare systems should be further evaluated.

Data from the general population support our findings.^31,32^ NT-proBNP and MR-proANP are independently associated with incident AF and improve performance of a model with clinical variables.^31^ Analysis of data from the LOOP trial demonstrates that people with elevated NT-proBNP benefit most from AF screening.^32^ Indeed, using an ILR to screen for AF in people aged ≥70 years prevented incident stroke or systemic embolism in people with NT-proBNP concentrations above the median, but not among people with lower NT-proBNP levels.^32^

AUROCs for models with MR-proANP or NT-proBNP were numerically higher in people without prior cardiac disease. This could be due to low prevalence of cardiac disease in the cohort. However, natriuretic peptides are elevated in people with cardiac disease which could influence performance in these subgroups. It is important to evaluate performance of MR-proANP and NT-proBNP in people with cardiac disease in future studies.

We determined the net reduction in people who would undergo PCM using a risk stratified approach to testing. The model with MR-proANP could reduce the number of people who would need PCM by 30%, compared to 27% for NT-proBNP and 20% for clinical variables alone. The estimates are based on 15% AFDAS threshold probability for PCM, which was selected using rates of AFDAS in the EMBRACE study and a meta-analysis of PCM after ischaemic stroke or TIA.^6,8^ Applying different AFDAS threshold probabilities would alter the net reduction in people who need PCM. However, models with MR-proANP or NT-proBNP would still reduce the number of people who need PCM by between 21% to 36% with AFDAS threshold probabilities of 10% to 20%, which would improve healthcare resource allocation for PCM.

### Strengths and Limitations

A major strength is the prospective, multi-centre nature of the BIOSIGNAL study, which includes people with a range of stroke subtypes rather than only including people with ESUS. This increases generalisability of our findings and is important given the STROKE-AF RCT demonstrates high rates of AFDAS in people with stroke attributed to small or large-artery disease.^33^ All patients had ≥3 days monitoring and the median duration is 9 days, which is important for AFDAS case ascertainment.

External validation was performed in the PRECISE study, which has a median duration of 28 days cardiac monitoring. PRECISE is a contemporaneous, prospective cohort study with robust AFDAS case ascertainment, which is important for external validation. We evaluated and compared the utility of MR-proANP and NT-proBNP alongside clinical variables to identify people who are unlikely to have AFDAS, which we believe is a novel approach. Moreover, we determined the net benefit of using the models to identify people who are unlikely to have AFDAS and guide cardiac monitoring approaches.

Our work also has several limitations. While the sample size is reasonable, it may have been insufficient to detect a statistically significant difference in performance of the model with MR-proANP compared to NT-proBNP. While the median duration of monitoring in the BIOSIGNAL cohort is 9 days, we cannot exclude some misclassification from missed cases of AFDAS. Finally, we did not assess ECG, echocardiogram or cerebral imaging parameters as these were not routinely available for the cohorts. We also wished to avoid developing approaches to guide cardiac monitoring that involve additional tests such as echocardiograms which are not routinely performed for all people after stroke.^10,34^

## Conclusions

MR-proANP and NT-proBNP can help classify people who are unlikely to have AFDAS. Measuring MR-proANP or NT-proBNP could reduce the number of people who need PCM after stroke and improve healthcare resource allocation.

## Data Availability

Ethical approval is not in place to share data from study participants.

## Acknowledgments

We acknowledge and thank all people who participated in the studies, and all members of the research teams at study sites.

---

**Supplemental Table 1.** Sensitivity Analyses Comparing Model Performance Across Subgroups.

| Subgroup | N (%) | AUROC<br>Clinical<br>Variables<br>(95% CI) | AUROC<br>Clinical Variables<br>+ log<sub>10</sub>MR-proANP<br>(95% CI) | p-value<br>(Clinical Variables<br>+ log<sub>10</sub>MR-proANP by<br>Subgroup) | AUROC<br>Clinical<br>Variables +<br>log<sub>10</sub>NT-proBNP<br>(95% CI) | p-value<br>(Clinical<br>Variables +<br>log<sub>10</sub>NT-proBNP<br>by Subgroup) | p-value<br>(Clinical Variables<br>+ log<sub>10</sub>MR-proANP<br>vs Clinical<br>Variables +<br>log<sub>10</sub>NT-proBNP) |
| --- | --- | --- | --- | --- | --- | --- | --- |
| Age > Median | 311<br>(50.1) | 0.57<br>(0.48-0.67) | 0.64<br>(0.56-0.72) | 0.40 | 0.62<br>(0.53-0.70) | 0.27 | 0.25 |
| Age ≤ Median | 310<br>(49.9) | 0.67<br>(0.54-0.80) | 0.72<br>(0.60-0.84) |  | 0.69<br>(0.55-0.83) |  | 0.27 |
| Female | 243<br>(39.1) | 0.67<br>(0.58-0.76) | 0.72<br>(0.63-0.80) | 0.86 | 0.70<br>(0.62-0.79) | 1.00 | 0.42 |
| Male | 378<br>(60.9) | 0.68<br>(0.60-0.77) | 0.72<br>(0.63-0.80) |  | 0.71<br>(0.63-0.80) |  | 0.40 |
| NIHSS ><br>Median | 279<br>(45) | 0.70<br>(0.62-0.79) | 0.74<br>(0.65-0.82) | 0.68 | 0.72<br>(0.64-0.80) | 0.59 | 0.34 |
| NIHSS ≤<br>Median | 341<br>(55) | 0.65<br>(0.56-0.74) | 0.70<br>(0.62-0.79) |  | 0.70<br>(0.61-0.78) |  | 0.63 |
| Cardiac Disease | 63<br>(10.2) | 0.60<br>(0.38-0.82) | 0.59<br>(0.39-0.80) | 0.18 | 0.59<br>(0.39-0.80) | 0.24 | 0.89 |
| No Cardiac<br>Disease | 557<br>(89.8) | 0.70<br>(0.63-0.76) | 0.74<br>(0.68-0.81) |  | 0.72<br>(0.66-0.79) |  | 0.11 |

**Supplemental Table 2.** Performance of the Models Across Durations of Cardiac Monitoring in the BIOSIGNAL Cohort.

|  | All Durations of Cardiac Monitoring | <3 Days of Cardiac Monitoring |
| --- | --- | --- |
| Model | AUROC<br>(95% CI) | AUROC<br>(95% CI) |
| Clinical Alone | 0.71<br>(0.68-0.74) | 0.70<br>(0.66-0.74) |
| Clinical and log<sub>10</sub>MR-proANP | 0.80<br>(0.77-0.82) | 0.82<br>(0.78-0.85) |
| Clinical and log<sub>10</sub>NT-proBNP | 0.78<br>(0.76-0.81) | 0.80<br>(0.77-0.83) |

**Supplemental Table 3.**
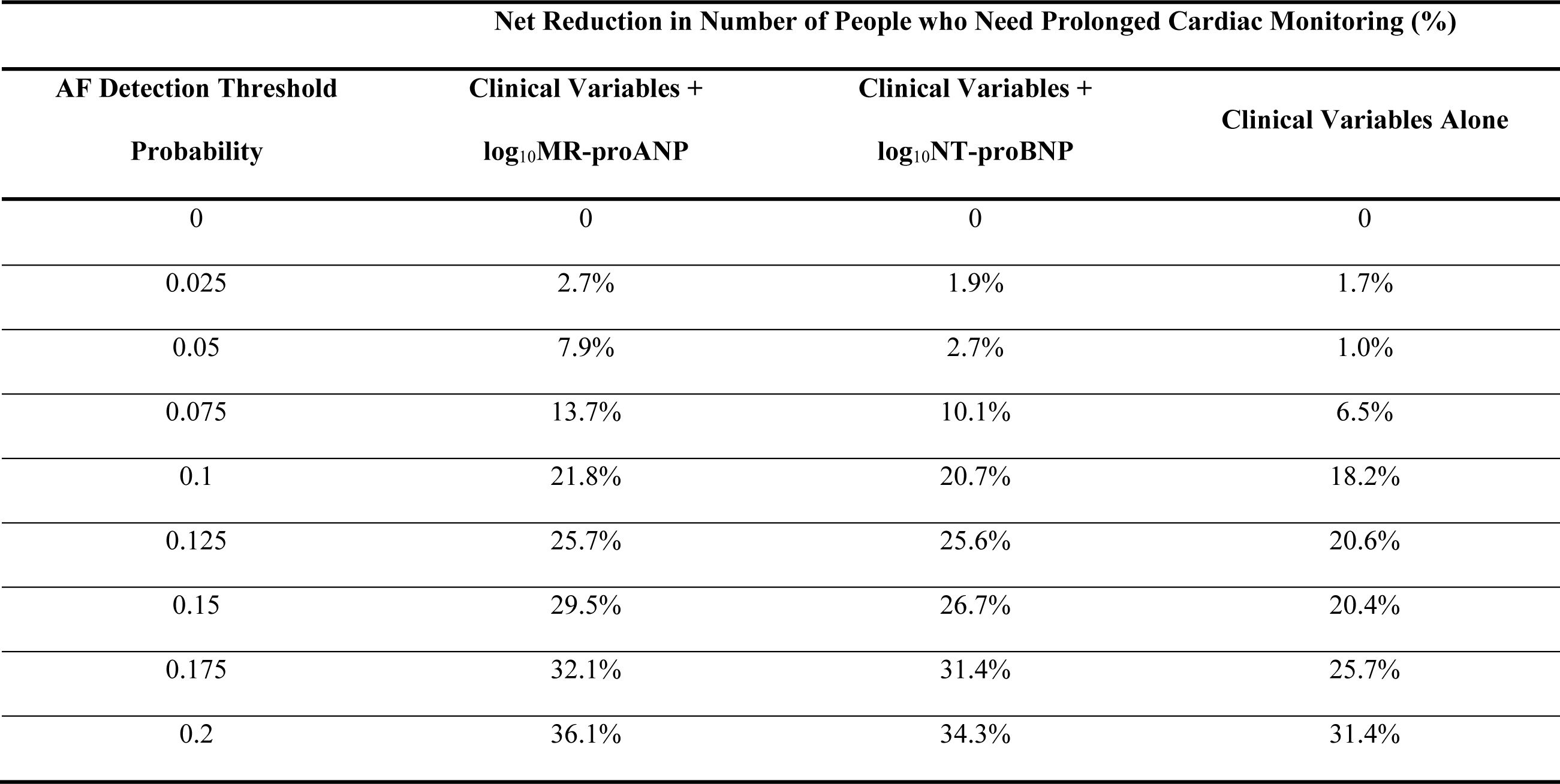
Decision Curve Analysis for Reduction in the Number of People who Need Prolonged Cardiac Monitoring.

| Net Reduction in Number of People who Need Prolonged Cardiac Monitoring (%) |  |  |  |
| --- | --- | --- | --- |
| AF Detection Threshold | Clinical Variables + | Clinical Variables + | Clinical Variables Alone |
| Probability | log<sub>10</sub>MR-proANP | log<sub>10</sub>NT-proBNP |  |
| 0 | 0 | 0 | 0 |
| 0.025 | 2.7% | 1.9% | 1.7% |
| 0.05 | 7.9% | 2.7% | 1.0% |
| 0.075 | 13.7% | 10.1% | 6.5% |
| 0.1 | 21.8% | 20.7% | 18.2% |
| 0.125 | 25.7% | 25.6% | 20.6% |
| 0.15 | 29.5% | 26.7% | 20.4% |
| 0.175 | 32.1% | 31.4% | 25.7% |
| 0.2 | 36.1% | 34.3% | 31.4% |

**Supplemental Figure 1.**
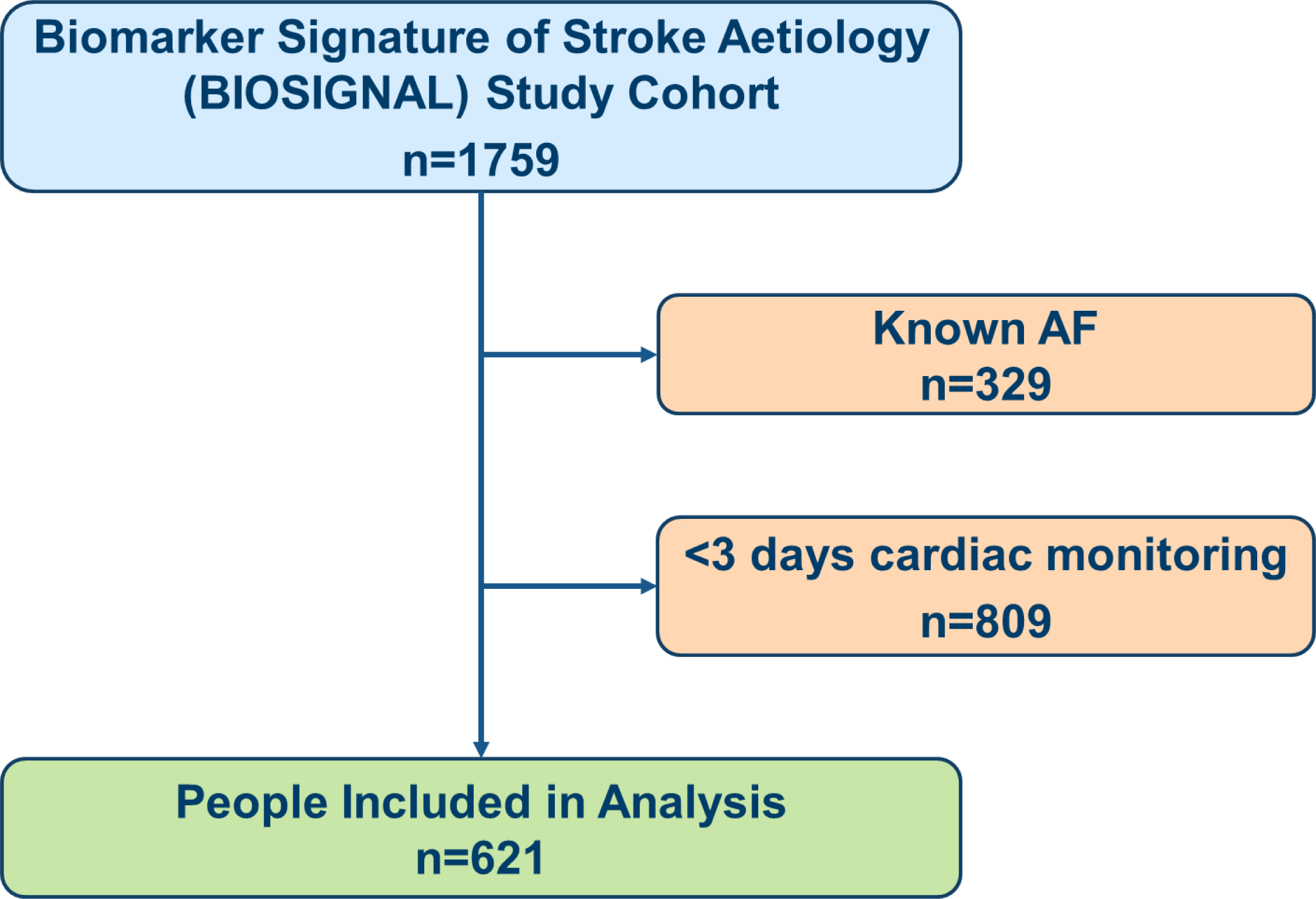
Overview of selection of people from the BIOSIGNAL study cohort for analysis.

## References

1. Ding M, Ebeling M, Ziegler L, Wennberg A, Modig K. Time trends in atrial fibrillation-related stroke during 2001-2020 in Sweden: a nationwide, observational study. Lancet Reg Health Eur. May 2023;28:100596. doi:10.1016/j.lanepe.2023.100596

2. Hart RG, Pearce LA, Aguilar MI. Meta-analysis: antithrombotic therapy to prevent stroke in patients who have nonvalvular atrial fibrillation. Ann Intern Med. Jun 19 2007;146(12):857–67. doi:10.7326/0003-4819-146-12-200706190-00007

3. Saxena R, Koudstaal PJ. Anticoagulants for preventing stroke in patients with nonrheumatic atrial fibrillation and a history of stroke or transient ischemic attack. Stroke. Jul 2004;35(7):1782–1783. doi:10.1161/01.Str.0000129900.70583.67

4. Hindricks G, Potpara T, Dagres N, et al. 2020 ESC Guidelines for the diagnosis and management of atrial fibrillation developed in collaboration with the European Association of Cardio-Thoracic Surgery (EACTS). Eur Heart J. Aug 29 2020;doi:10.1093/eurheartj/ehaa612

5. Grond M, Jauss M, Hamann G, et al. Improved detection of silent atrial fibrillation using 72-hour Holter ECG in patients with ischemic stroke: a prospective multicenter cohort study. Stroke. Dec 2013;44(12):3357–64. doi:10.1161/STROKEAHA.113.001884

6. Gladstone DJ, Spring M, Dorian P, et al. Atrial fibrillation in patients with cryptogenic stroke. N Engl J Med. Jun 26 2014;370(26):2467–77. doi:10.1056/NEJMoa1311376

7. Israel C, Kitsiou A, Kalyani M, et al. Detection of atrial fibrillation in patients with embolic stroke of undetermined source by prolonged monitoring with implantable loop recorders. Thromb Haemostasis. Oct 2017;117(10):1962–1969. doi:10.1160/Th17-02-0072

8. Tsivgoulis G, Triantafyllou S, Palaiodimou L, et al. Prolonged Cardiac Monitoring and Stroke Recurrence: A Meta-analysis. Neurology. May 10 2022;98(19):e1942–e1952. doi:10.1212/WNL.0000000000200227

9. Sposato LA, Cipriano LE, Saposnik G, Vargas ER, Riccio PM, Hachinski V. Diagnosis of atrial fibrillation after stroke and transient ischaemic attack: a systematic review and meta-analysis. Lancet Neurol. Apr 2015;14(4):377–387. doi:10.1016/S1474-4422(15)70027-X

10. Kleindorfer DO, Towfighi A, Chaturvedi S, et al. 2021 Guideline for the Prevention of Stroke in Patients With Stroke and Transient Ischemic Attack: A Guideline From the American Heart Association/American Stroke Association. Stroke. Jul 2021;52(7):e364–e467. doi:10.1161/STR.0000000000000375

11. Rubiera M, Aires A, Antonenko K, et al. European Stroke Organisation (ESO) guideline on screening for subclinical atrial fibrillation after stroke or transient ischaemic attack of undetermined origin. European Stroke Journal. 2022;doi:10.1177/23969873221099478

12. Geraghty O, Korompoki E, Filippidis FT, Rudd A, Veltkamp R. Cardiac diagnostic work-up for atrial fibrillation after transient ischaemic attacks in England and Wales: results from a cross-sectional survey. Bmj Open. 2016;6(11) doi: 10.1136/bmjopen-2016-012714

13. Kwong C, Ling AY, Crawford MH, Zhao SX, Shah NH. A Clinical Score for Predicting Atrial Fibrillation in Patients with Cryptogenic Stroke or Transient Ischemic Attack. Cardiology. 2017;138(3):133–140. doi:10.1159/000476030

14. Uphaus T, Weber-Kruger M, Grond M, et al. Development and validation of a score to detect paroxysmal atrial fibrillation after stroke. Neurology. Jan 8 2019;92(2):e115–e124. doi:10.1212/WNL.0000000000006727

15. Ntaios G, Perlepe K, Lambrou D, et al. External Performance of the HAVOC Score for the Prediction of New Incident Atrial Fibrillation. Stroke. Feb 2020;51(2):457–461. doi:10.1161/STROKEAHA.119.027990

16. Ntaios G, Perlepe K, Lambrou D, et al. Identification of patients with embolic stroke of undetermined source and low risk of new incident atrial fibrillation: The AF-ESUS score. Int J Stroke. Jan 2021;16(1):29–38. doi:10.1177/1747493020925281

17. Kitsiou A, Sagris D, Schabitz WR, Ntaios G. Validation of the AF-ESUS score to identify patients with embolic stroke of undetermined source and low risk of device-detected atrial fibrillation. Eur J Intern Med. Jul 2021;89:135–136. doi:10.1016/j.ejim.2021.04.003

18. Koziel M, Potpara TS, Lip GYH. Using Blood Biomarkers to Identify Atrial Fibrillation-Related Stroke. Stroke. Aug 2019;50(8):1956–1957. doi:10.1161/STROKEAHA.119.026185

19. De Marchis GM, Schneider J, Weck A, et al. Midregional proatrial natriuretic peptide improves risk stratification after ischemic stroke. Neurology. Feb 6 2018;90(6):e455–e465. doi:10.1212/WNL.0000000000004922

20. Schweizer J, Arnold M, Konig IR, et al. Measurement of Midregional Pro-Atrial Natriuretic Peptide to Discover Atrial Fibrillation in Patients With Ischemic Stroke. J Am Coll Cardiol. Apr 12 2022;79(14):1369–1381. doi:10.1016/j.jacc.2022.01.042

21. Wasser K, Weber-Kruger M, Groschel S, et al. Brain Natriuretic Peptide and Discovery of Atrial Fibrillation After Stroke: A Subanalysis of the Find-AFRANDOMISED Trial. Stroke. Feb 2020;51(2):395–401. doi:10.1161/STROKEAHA.119.026496

22. Morgenthaler NG, Struck J, Thomas B, Bergmann A. Immunoluminometric assay for the midregion of pro-atrial natriuretic peptide in human plasma. Clin Chem. Jan 2004;50(1):234–6. doi:10.1373/clinchem.2003.021204

23. Schnabel RB, Haeusler KG, Healey JS, et al. Searching for Atrial Fibrillation Poststroke: A White Paper of the AF-SCREEN International Collaboration. Circulation. Nov 26 2019;140(22):1834–1850. doi:10.1161/CIRCULATIONAHA.119.040267

24. Robin X, Turck N, Hainard A, et al. pROC: an open-source package for R and S+ to analyze and compare ROC curves. BMC Bioinformatics. Mar 17 2011;12:77. doi:10.1186/1471-2105-12-77

25. DeLong ER, DeLong DM, Clarke-Pearson DL. Comparing the areas under two or more correlated receiver operating characteristic curves: a nonparametric approach. Biometrics. Sep 1988;44(3):837–45.

26. Cameron AC, Katsas G, Arnold M, et al. Preventing Recurrent Cardioembolic Stroke: Right Approach, Right Patient (PRECISE) Study Protocol. Cerebrovasc Dis. Aug 29 2022:1–7. doi:10.1159/000525918

27. Vickers AJ, van Calster B, Steyerberg EW. A simple, step-by-step guide to interpreting decision curve analysis. Diagn Progn Res. 2019;3:18. doi:10.1186/s41512-019-0064-7

28. Vickers AJ, Van Calster B, Steyerberg EW. Net benefit approaches to the evaluation of prediction models, molecular markers, and diagnostic tests. BMJ. Jan 25 2016;352 doi: 10.1136/bmj.i6

29. Kishore AK, Hossain MJ, Cameron A, Dawson J, Vail A, Smith CJ. Use of risk scores for predicting new atrial fibrillation after ischemic stroke or transient ischemic attack-A systematic review. International Journal of Stroke. Jul 2022;17(6):608–617. doi: 10.1177/17474930211045880

30. McDonagh TA, Metra M, Adamo M, et al. 2021 ESC Guidelines for the diagnosis and treatment of acute and chronic heart failure Developed by the Task Force for the diagnosis and treatment of acute and chronic heart failure of the European Society of Cardiology (ESC) With the special contribution of the Heart Failure Association (HFA) of the ESC. Eur J Heart Fail. Jan 2022;24(1):4–131. doi:10.1002/ejhf.2333

31. Schnabel RB, Wild PS, Wilde S, et al. Multiple biomarkers and atrial fibrillation in the general population. PLoS One. 2014;9(11):e112486. doi:10.1371/journal.pone.0112486

32. Xing LY, Diederichsen SZ, Hojberg S, et al. Effects of Systematic Atrial Fibrillation Screening According to N-Terminal Pro-B-Type Natriuretic Peptide: a Secondary Analysis of the Randomized LOOP Study. Circulation. Jun 2023;147(24):1788–1797 doi:10.1161/CIRCULATIONAHA.123.064361

33. Bernstein RA, Kamel H, Granger CB, et al. Effect of Long-term Continuous Cardiac Monitoring vs Usual Care on Detection of Atrial Fibrillation in Patients With Stroke Attributed to Large- or Small-Vessel Disease: The STROKE-AF Randomized Clinical Trial. JAMA. Jun 2021;325(21):2169–2177. doi:10.1001/jama.2021.6470

34. European Stroke Organisation Executive Committee. Guidelines for management of ischaemic stroke and transient ischaemic attack 2008. Cerebrovasc Dis. 2008;25(5):457–507. doi:10.1159/000131083

